# Age-specific case data reveal varying dengue transmission intensity in US states and territories

**DOI:** 10.1101/2023.02.07.23285569

**Authors:** Sarah Kada, Gabriela Paz-Bailey, Laura Adams, Michael A. Johansson

## Abstract

Dengue viruses (DENV) are endemic in the US territories of Puerto Rico, American Samoa, and the US Virgin Islands, with focal outbreaks also reported in the states of Florida and Hawaii. However, little is known about the intensity of dengue virus transmission over time and how dengue viruses have shaped the level of immunity in these populations, despite the importance of understanding how and why levels of immunity against dengue may change over time. These changes need to be considered when responding to future outbreaks and enacting dengue management strategies, such as guiding vaccine deployment. We used catalytic models fitted to case surveillance data stratified by age from the ArboNET national arboviral surveillance system to reconstruct the history of recent dengue virus transmission in Puerto Rico, American Samoa, US Virgin Islands, Florida, Hawaii, and Guam. We estimated average annual transmission intensity (i.e., force of infection) of DENV between 2010 and 2019 and the level of seroprevalence by age group in each population. We compared models and found that treating all reported cases as secondary infections generally fit the surveillance data better than models considering cases as primary infections. Using the secondary case model, we found that force of infection was highly heterogeneous between jurisdictions and over time within jurisdictions, ranging from 0.00003 (95% CI: 0.00002–0.0004) in Florida to 0.08 (95% CI: 0.044–0.14) in American Samoa during the 2010–2019 period. For early 2020, we estimated that seropositivity in 10 year-olds ranged from 0.09% (0.02%–0.54%) in Florida to 56.3% (43.7%–69.3%) in American Samoa. In the absence of serological data, age-specific case notification data collected through routine surveillance combined with mathematical modeling are powerful tools to monitor arbovirus circulation, estimate the level of population immunity, and design dengue management strategies.

**Author Summary:** Viruses transmitted by *Aedes* mosquitoes are a substantial public health burden globally. In the US, the increasing number of outbreaks in recent decades and the co-circulation of the four dengue viruses present a risk of experiencing sequential infections, which can result in more severe disease. However, reported cases only represent a proportion of all infections, so the intensity of dengue circulation in US jurisdictions and the level of population immunity are largely unknown. Combining dengue surveillance data stratified by age with mathematical models enables direct estimation of transmission intensity and previous exposure. We found that dengue virus transmission intensity was highest in American Samoa and Puerto Rico. The US Virgin Islands also showed patterns of endemic transmission, while Hawaii and Guam present more sporadic outbreak profiles. These estimates can help improve risk assessment in these locations and guide interventions to the populations who most need them.

## Introduction

Dengue viruses (DENV) re-emerged in the Americas beginning in the 1970s, with the viruses rapidly expanding their range and associated burden of dengue infections [1–3]. However, this expansion has not been uniform. The four antigenically distinct dengue virus serotypes (DENV-1 to DENV-4) emerged at different times with affected areas experiencing different levels of transmission intensity, from sporadic outbreaks to hyperendemic, year-round circulation [4–6].

The varied history of DENV transmission in the Americas directly shapes the current risk. Infection with one serotype generally provides long-term immunity to that serotype and short-term immunity to all serotypes. Secondary exposure to a different serotype from the primary infection has been associated with increased risk of severe dengue [7]. Population immunity is therefore modulated by the circulation of the four serotypes and the intensity of transmission of those viruses.

As population immunity and transmission intensity are intrinsically linked, it is challenging to estimate either of those components directly. Existing immunity can be partially measured with age-specific serological surveys, but these are logistically and financially challenging to implement, provide only a snapshot of previous exposure, and differentiating specific serotypes and the sequence of prior serotype exposures is very difficult. Transmission intensity can be characterized as the force of infection (FOI), the per capita rate at which susceptible individuals become infected by an infectious disease. A class of models called catalytic models can estimate the FOI, providing a measure of changes in exposure to diseases. However, direct estimation of the FOI is also challenging because population susceptibility is generally not known, many dengue infections are inapparent or unreported, and the proportion of individuals experiencing inapparent disease is dependent on prior infection [8–11]. Estimating the FOI therefore requires accounting for both underreporting and historical transmission intensity.

To help meet this challenge, age-stratified case data collected through passive surveillance can be used to gather insight into both existing immunity and transmission intensity [12]. Due to every age group having different exposure periods, incidence across each age group each year reflects both the age-group specific immunity and the yearly transmission intensity, enabling estimation of yearly transmission intensity for both current and past years [13]. Catalytic models that use age-specific case data can thus provide valuable insights into the dynamics of arboviruses [12,14,15] by quantifying infection burden in settings where seroprevalence data are unavailable or unrepresentative of the population.

Here, we analyze reported dengue case data from six US jurisdictions with outbreaks reported since dengue became a nationally reportable disease in 2010: Puerto Rico, American Samoa, US Virgin Islands, Florida, Hawaii and Guam. These locations represent diverse historical trends in reported case data, from long-term hyper-endemicity in Puerto Rico to no reported dengue in decades prior to an outbreak in 2019 in Guam (Figure 1). Building on previous work by Rodriguez-Barraquer et al. [12], we developed a generalized model that accounts for variability in transmission intensity and reporting over time and estimates the FOI and age-specific immunity over time. Accounting for yearly variability in FOI and reporting provides critical insights into dengue epidemiology in a wide variety of settings, from hyperendemic to sporadic outbreaks, and can be used to better assess dengue burden in the US and other settings, contribute to epidemic risk assessment, and guide implementation and evaluation of control strategies such as vaccination.

**Figure 1.**
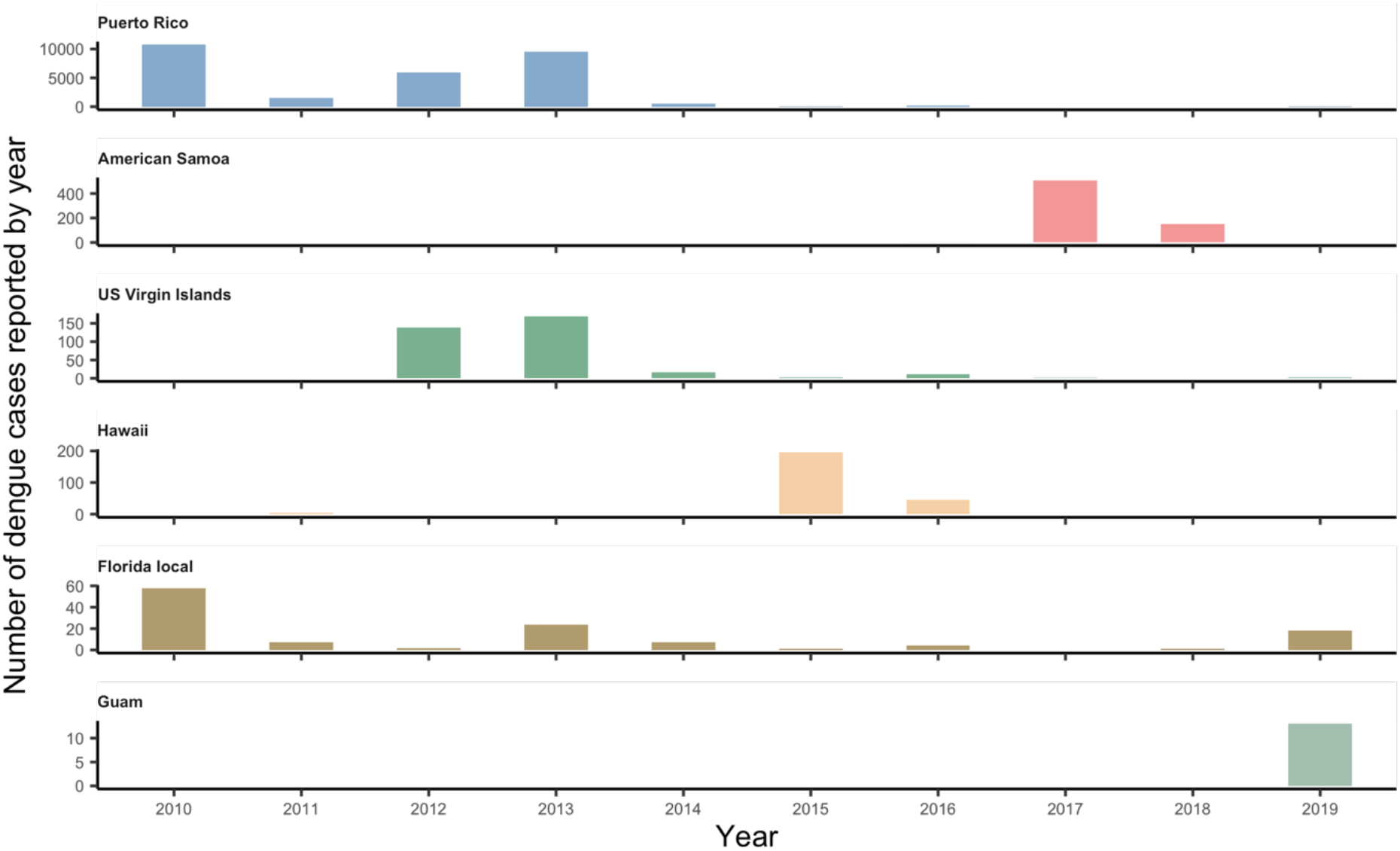
Confirmed and probable dengue cases reported to ArboNET by state or territory, 2010–2019. Included cases had no travel history and met laboratory criteria for diagnosis. Different y-axis scales were used for each jurisdiction.

## Methods

### Data sources

Dengue became a nationally notifiable disease in January 2010 and we analyzed confirmed and probable dengue cases reported to ArboNET for the years 2010–2019 for six jurisdictions: Puerto Rico, American Samoa, US Virgin Islands, Hawaii, Florida and Guam. Texas was excluded due to low case counts. ArboNET includes information about place of likely acquisition (locally acquired or travel-associated) and disease severity. We analyzed locally acquired cases only, except Florida, where we analyzed both categories. For all locations, we combined severe and non-severe cases. In Puerto Rico, where the highest number of severe cases was reported, we also analyzed severe cases (including dengue hemorrhagic fever and dengue shock syndrome [16,17]) on their own. Age distribution data for each population was obtained from the 2010 census data provided by the United States Census Bureau (census.gov), except for Florida, Hawaii and Puerto Rico where in addition to the 2010 census data, population estimates were available for years 2011–2019. The ArboNET surveillance data is available to researchers upon request.

### Estimating the force of infection

We used a Bayesian catalytic model extending one developed by Rodriguez-Barraquer et al., [12] to estimate the time-dependent force of infection (FOI(t)) of all circulating serotypes and derive age-specific seroprevalence estimates. Building on the original model, we (i) added random effects for both reporting and FOI probabilities to allow a long-term mean with yearly variability for each of these components. We also (ii) implemented separate models for different reporting possibilities (cases reported are primary DENV infections, secondary infections or a combination of both) and (iii) accounted for potential infections in infants that appear more like secondary infections due to possible maternal antibody transfer. Finally, we (iv) generated a full suite of retrospective FOI estimates (prior to 2010, i.e., when ArboNET data became available).

#### Reporting probability

Many DENV infections are mild or asymptomatic, and do not result in a visit to a clinic or being reported as a case [8,9,18]. Building on previous estimates, we set a prior for the probability of reporting a case using a logit-normal distribution (mean = -2.2, β = 0.7) for the long-term mean, which translates into a mean value of approximately 10% with a 97.5% bound of 30%. Multiple factors may lead to year-to-year fluctuation of reporting probabilities; the presence of other arboviruses infections may influence dengue reporting and testing, and, during epidemic years, doctors may be more likely to recognize and test dengue cases [19–22]. To allow some variability in reporting probabilities, we let it vary between years and partitioned the variance of the reporting prior between uncertainty in the mean and uncertainty in year-to-year variability (see S1).

The probability of identifying and reporting dengue cases is also dependent on case severity. In one cohort study, the proportion of inapparent versus symptomatic case was similar between primary and secondary cases, with age, year, and time interval between consecutive infections more likely to affect symptomatic status [23]. Additionally, laboratory confirmation generally relies on virological testing that cannot differentiate primary and secondary infections, so the proportion of cases reported to ArboNET that are primary or secondary is not known. We therefore compared three different models considering either that all reported cases are secondary infections (Model S), that reported cases are a combination of primary and secondary infections (Model PS), and that all reported cases are primary infections (Model P, see Supporting information S1).

Severe dengue cases, which are predominantly secondary infections, are also reported to ArboNET. Because substantial numbers of severe cases were reported in Puerto Rico during the study time period, we performed a secondary analysis for Puerto Rico limiting the included cases to severe dengue only. While severe cases are more likely to be reported than non-severe cases, severe cases represent only a small proportion of secondary infections. In previous cohort studies, the estimated probability of a secondary infection resulting in a reported severe case was lower than the reporting probability of a secondary infection resulting in any reported dengue case (dengue fever or severe dengue), below 5% (less than 1% [23] and 3% [24]). Based on those studies, we used a lower prior for the reporting of severe cases, setting a mean of 1% of all primary or secondary (depending on the model) infections being reported as severe cases and a 97.5% bound of 5%.

#### FOI

The FOI is likely to vary over time depending on the circulating dengue serotypes, serotype introduction or re-introduction, and population immunity, but few catalytic models account for this fluctuation [25]. Similar to the reporting probabilities (described above), we use a logit-normal prior for long-term average FOI and partitioned the prior variance between uncertainty in the mean and uncertainty in the year-to-year variability. We estimated a specific FOI prior for each location, using the average age of infection, life expectancy, and the mean reporting prior (see Supporting information S1, Table S1).

#### Infant cases

The presence of maternal antibodies may interfere and potentially predispose some infants to more severe disease, such that infant infections may be more severe and more likely to be reported than expected for a true primary infection [26]. To account for this, we added a proportion parameter, *α*, that sets the initial previous exposure for infants to a proportion of the FOI in the previous year rather than zero (see S1, Figure S13).

### Model comparison

Models were compared using leave-one-out (loo) Pareto smoothed importance sampling [27]. Lower loo scores indicate better cross-validation model fit. We also collected yearly log-likelihood estimates to assess how our S, P and PS models fit to the reported case data. For each parameter we extracted the posterior medians and 95% Bayesian credible intervals (hereafter 95% CI). Models were run using Bayesian Markov Chain Monte Carlo sampling with a No-U-Turn sampler using the RStan package [28] in R (version 4.1.2, [29]). Detailed information on the methods used can be found in Supporting Information S1.

## Results

Dengue circulation was highly heterogeneous during 2010–2019 across the six jurisdictions (Figure 1). The age distribution of cases also varied markedly, with the most commonly reported age group being 10–14 years in the US Virgin Islands and Guam to 60–64 years for locally acquired cases in Florida (Figure 2). If all age groups had the same risk of infection, the age distribution of cases would mirror the population age distribution, e.g., the largest age group is more likely to have the highest number of reported cases. However, immunity from prior infection, especially in endemic areas, shifts the age of cases to predominantly younger age groups (e.g., Puerto Rico, American Samoa).

**Figure 2.**
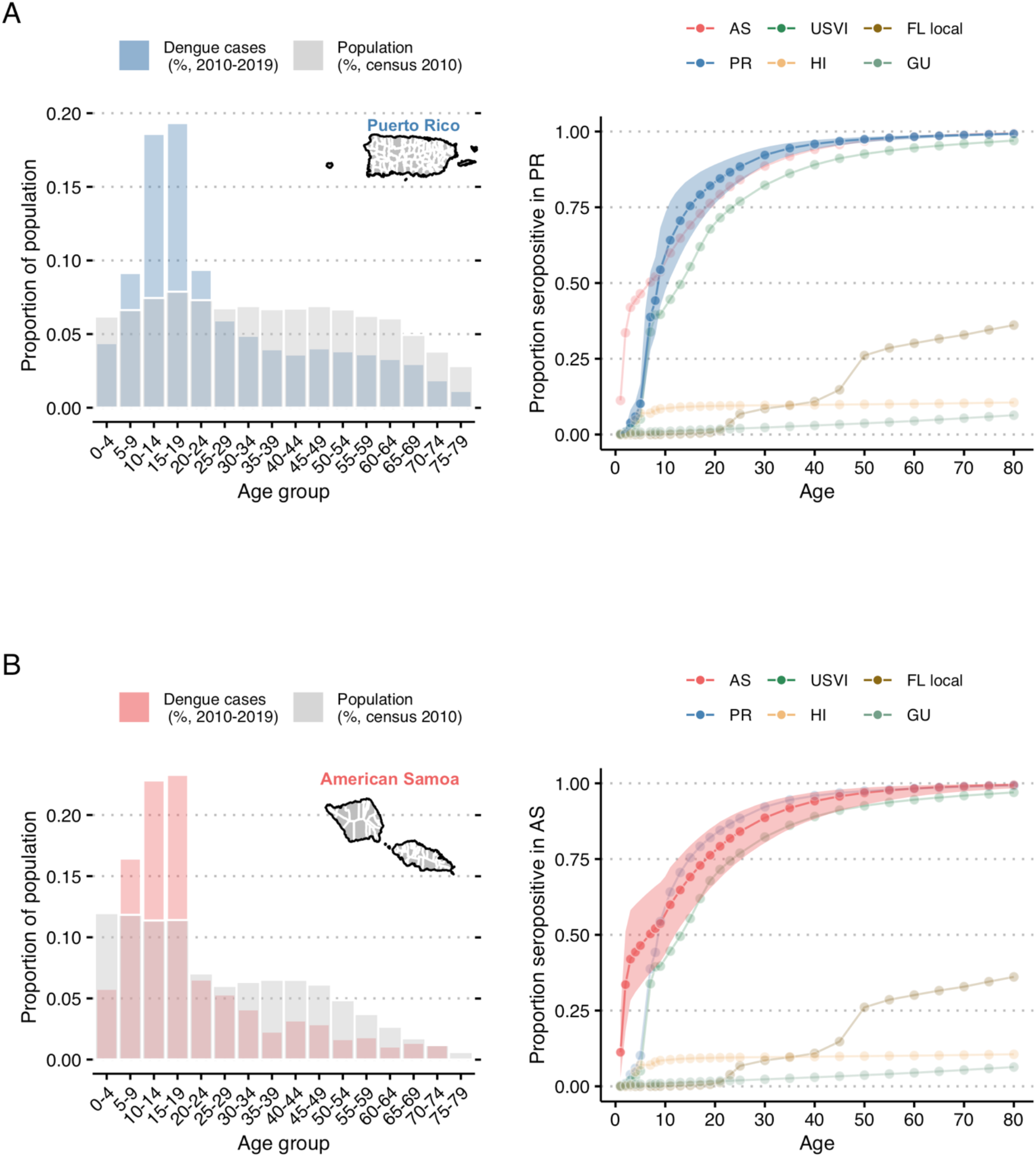
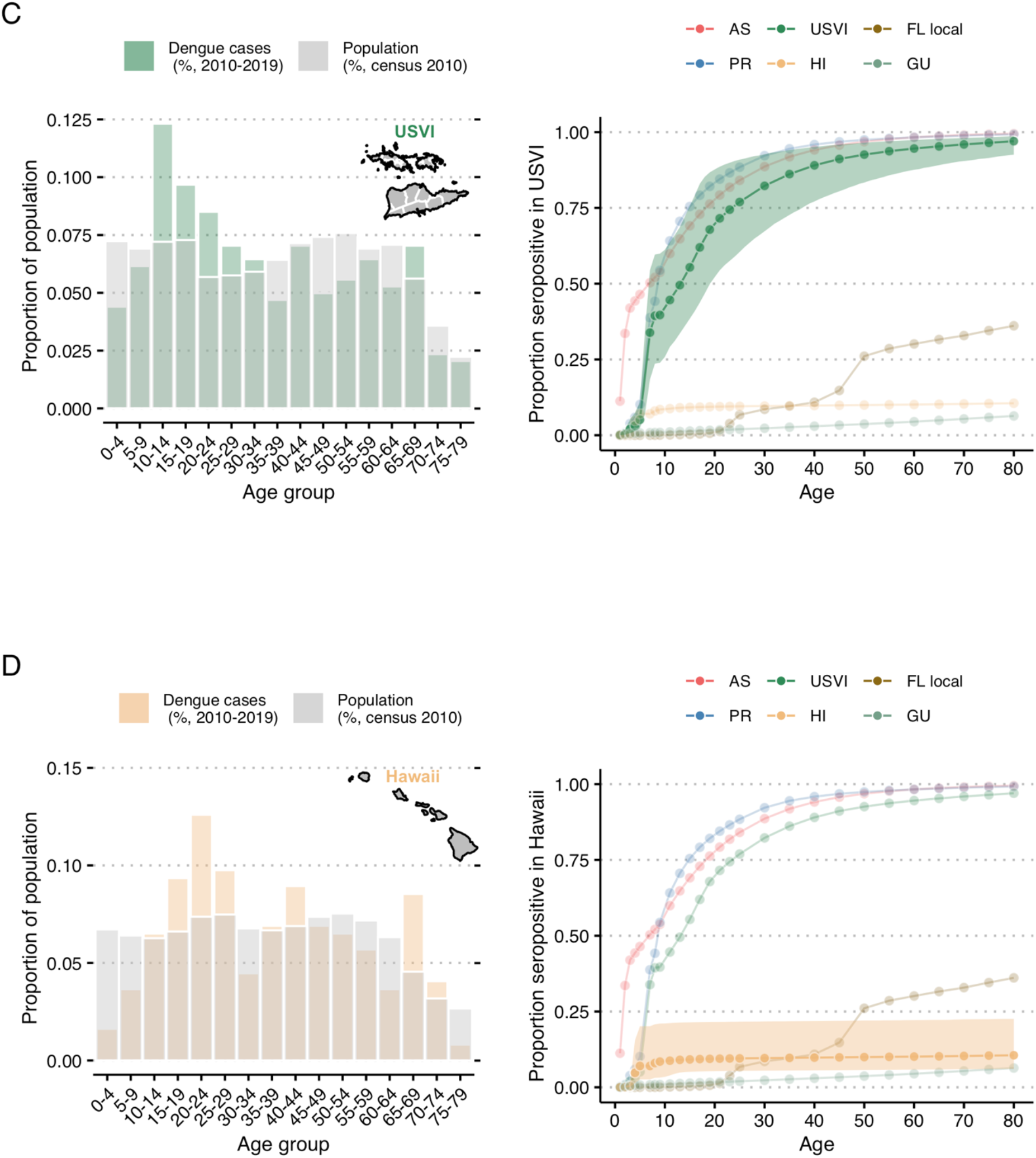
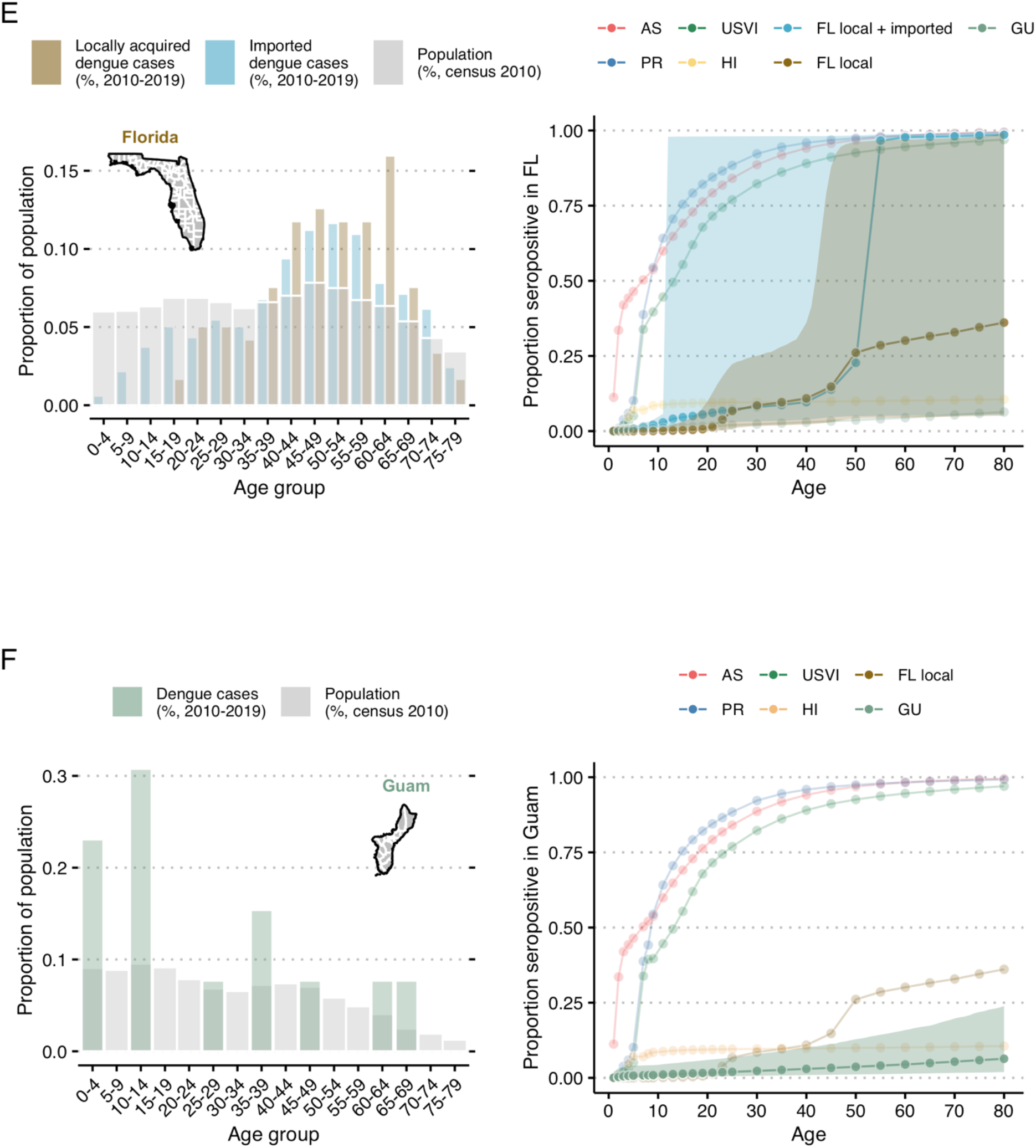
Age distribution of reported dengue cases and 2019 seroprevalence estimates by age group for each US state and territory assessed. Age distribution of all locally reported, confirmed and probable dengue cases in Puerto Rico (PR, A), American Samoa (AS, B), US Virgin Islands (USVI, C), Hawaii (HI, D), Florida (FL, E) and Guam (GU, F) in 2010–2019 (colored bar) and age distribution of the population in these respective territories (grey bars, 2010 census) (left panel) and age distribution of 2019 seroprevalence estimate obtained from the fit of model S (reported cases are secondary infections only), with corresponding 95% confidence intervals (shaded area, right panel). Florida (E) contains the additional seroprevalence estimates from all cases (imported and local). Detailed tables on years with reported dengue cases are available in S1. Y-axis scale on the left panels differ for each location.

We fitted models to each time series of age-specific case data under three different assumptions: (i) that all reported cases are primary infections (Model P), (ii) that all reported cases are secondary infections (Model S), and (iii) that reported cases include both primary and secondary cases (Model PS). In Hawaii and Guam, Model P and Model PS had significantly lower loo error than Model S (Figure 3). The fit of Model S was only significantly better for local cases in Florida (i.e., excluding travel-associated cases) though also had lower mean error for the US Virgin Islands and severe cases in Puerto Rico.

**Figure 3.**
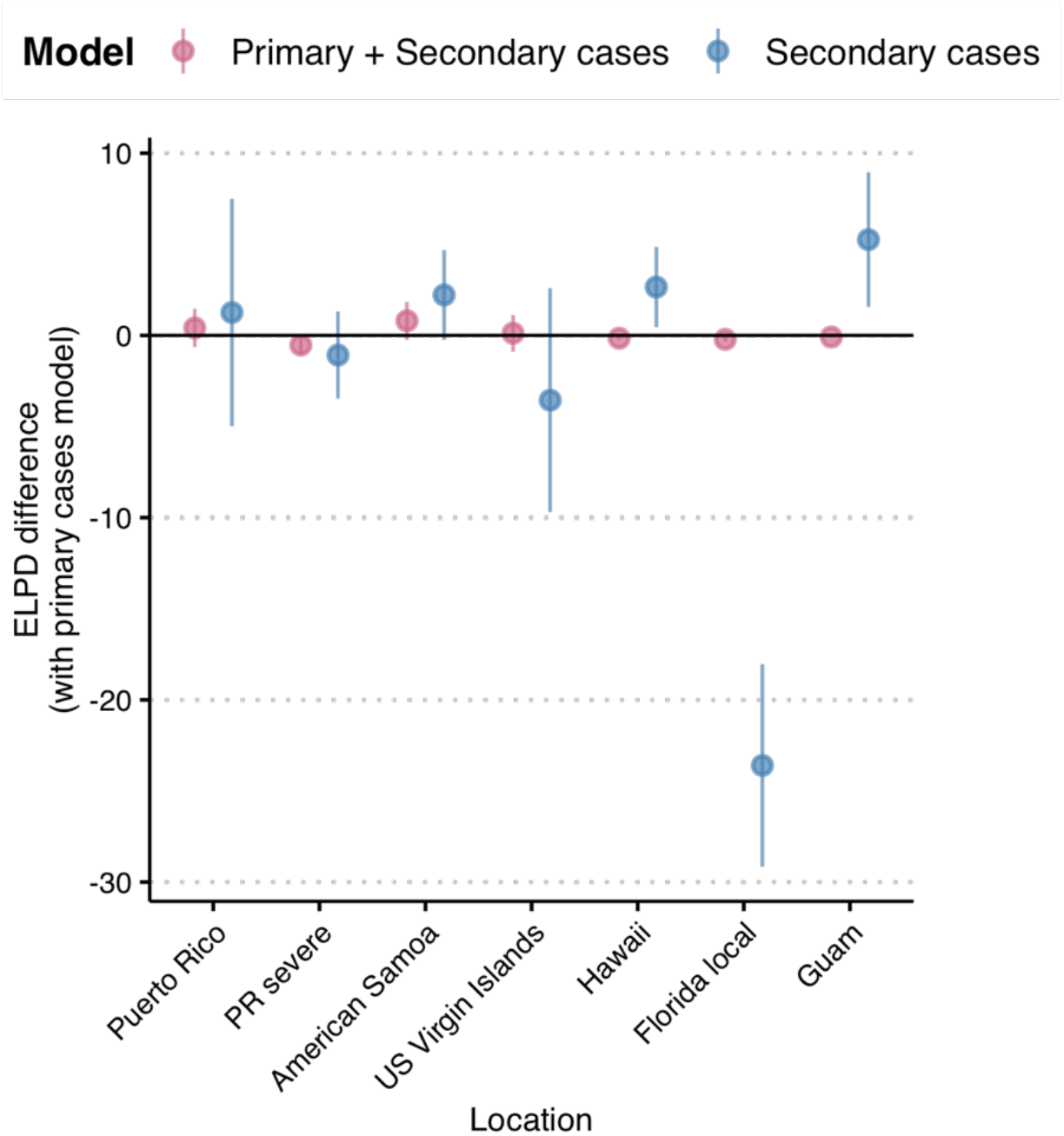
Leave-one-out cross validation model comparison using the estimated difference in ELPD (expected log pointwise predictive probabilities). ELPD is a measure of out-of-sample predictive accuracy, as estimated by the Bayesian leave-one-out cross validation (LOO). We compared model P (cases as primary) to models PS and S (primary and secondary cases and secondary cases only). Models under the horizontal bar at 0 fit the data better than model P (reference). Vertical bars represent the corresponding standard error of the difference in ELPD.

Comparing the different models across years, age groups and locations, we found mixed evidence about relative fit. For Puerto Rico, log likelihoods indicated that Model S tended to have a better fit in years with more reported cases and sometimes a worse fit in years and age groups with fewer reported cases (S1, Figures S5-S6). Model S also appeared to fit the distribution of cases across younger age groups more accurately across most jurisdictions (S1, Figures S7 to S13). Models P and PS both tend to estimate that a larger number of cases should be reported in the youngest age group (0-4 years old). Model S qualitatively better matches the distinct pattern of dengue cases in children across endemic areas. Because of this distinct advantage and the finding that fit for Model S was only inferior for Hawaii and Guam (where there was the least evidence of sustained transmission) (S1, Figures S7 to S13), we used Model S for subsequent analyses.

Of the six locations analyzed, Puerto Rico had the second highest long-term mean estimated FOI, an estimated 1.1% (95% CI: 0.9–1.3%) per year with substantial year-to-year variation including higher FOI estimates in the early 1990s and 2000s (prior to the dataset analyzed here, Figure 4). The estimated probability of a secondary infection resulting in a reported dengue case was 2.1% (95% CI: 1.4–3.3%). The estimated probability of a secondary infection resulting in a reported severe dengue case was 0.1% (95% CI: 0.1–0.3%). Reporting was estimated to be slightly higher in years when more cases were reported (Figure 5). For 2019, we estimated seroprevalence in 5-year-old children to be 10.1% (95% CI: 6.1–16.9%), increasing to 94.5% (95% CI: 92–96.2%) by age 35 (Figure 2A). FOI and seroprevalence estimates using only severe cases were similar to those using all cases in Puerto Rico, although we observe a lower seroprevalence for 2019 in younger age groups and higher in older age groups (Figure 6).

**Figure 4.**
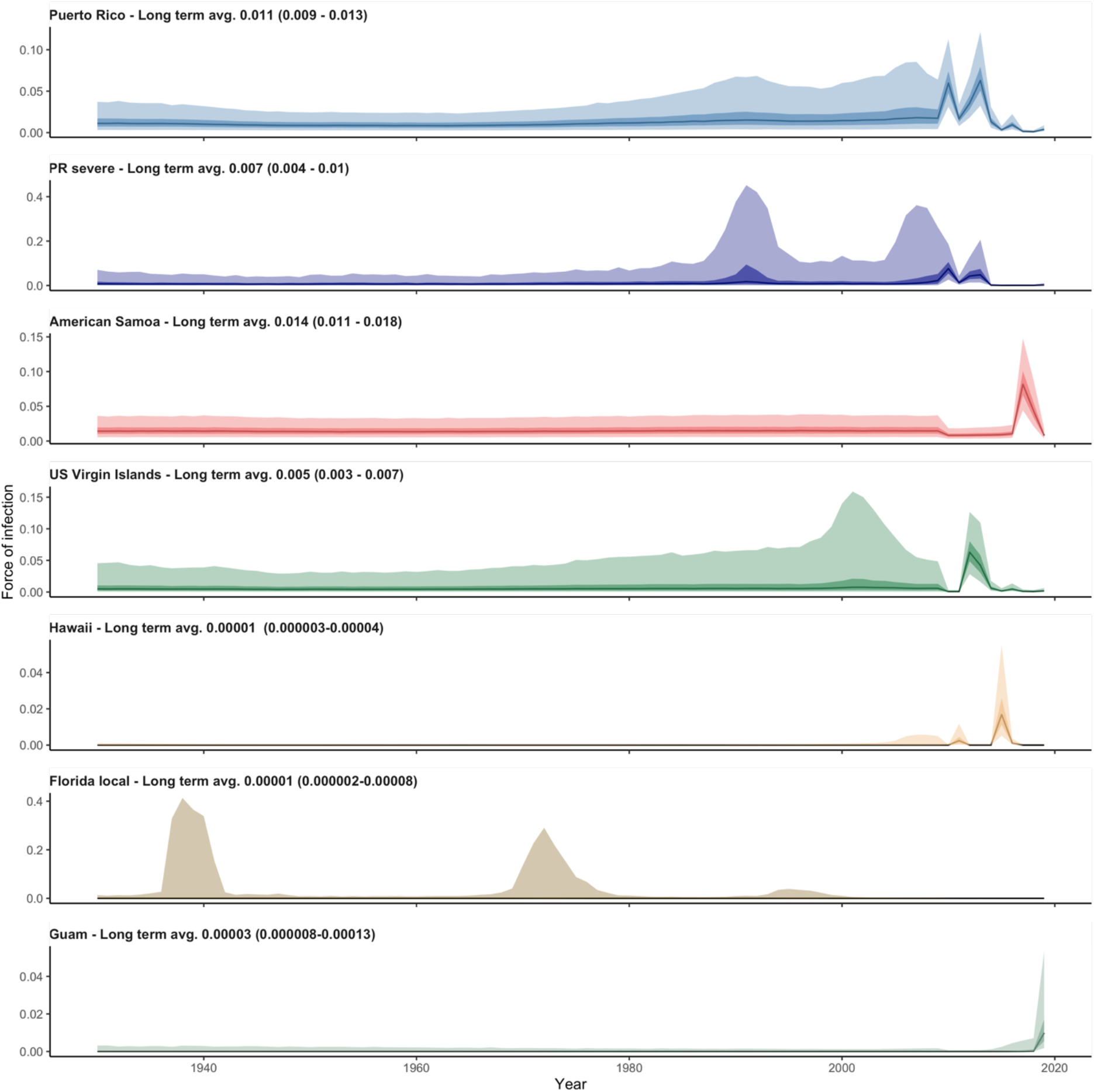
Time-varying estimates of the force of infection (FOI) in Puerto Rico (all cases) and severe cases only (PR severe), American Samoa, US Virgin Islands, Hawaii, Florida local cases and Guam in 2010–2019 from our model S (cases as secondary infections). The numbers in the upper left-hand corner of each panel shows long term FOI estimates for each location. Dark and light shaded areas represent respectively, 50% and 95% confidence intervals. Different y-axis scales were used for each jurisdiction.

**Figure 5.**
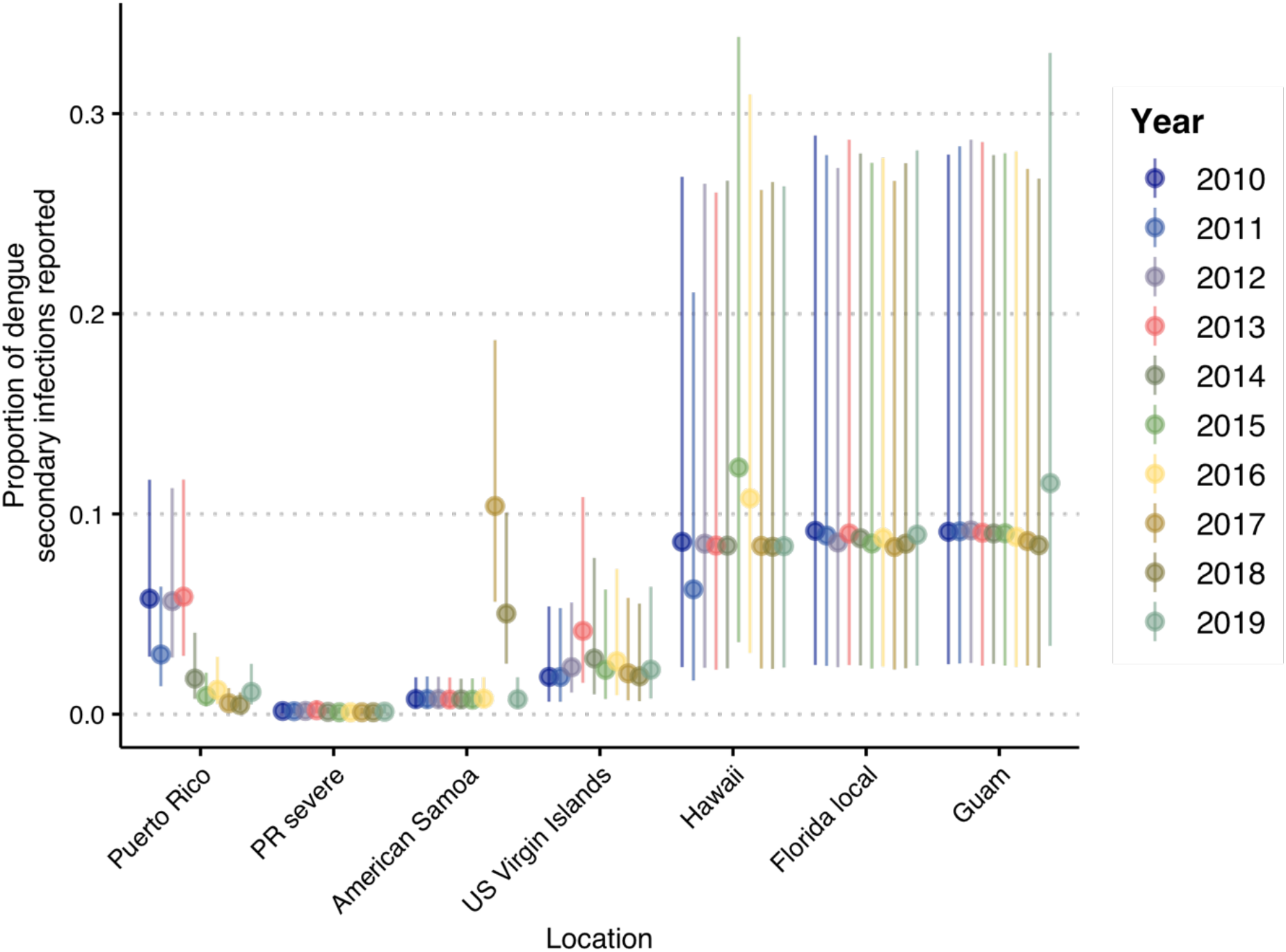
Estimates of the yearly probability of reporting cases in Puerto Rico (all cases) and severe cases only (PR severe), American Samoa, US Virgin Islands, Hawaii, Florida and Guam in 2010–2019 from our S model (cases are considered secondary infections). Vertical bars represent 95% confidence intervals. For the severe cases, it is the proportion of all secondary infections that are both severe and reported.

**Figure 6.**
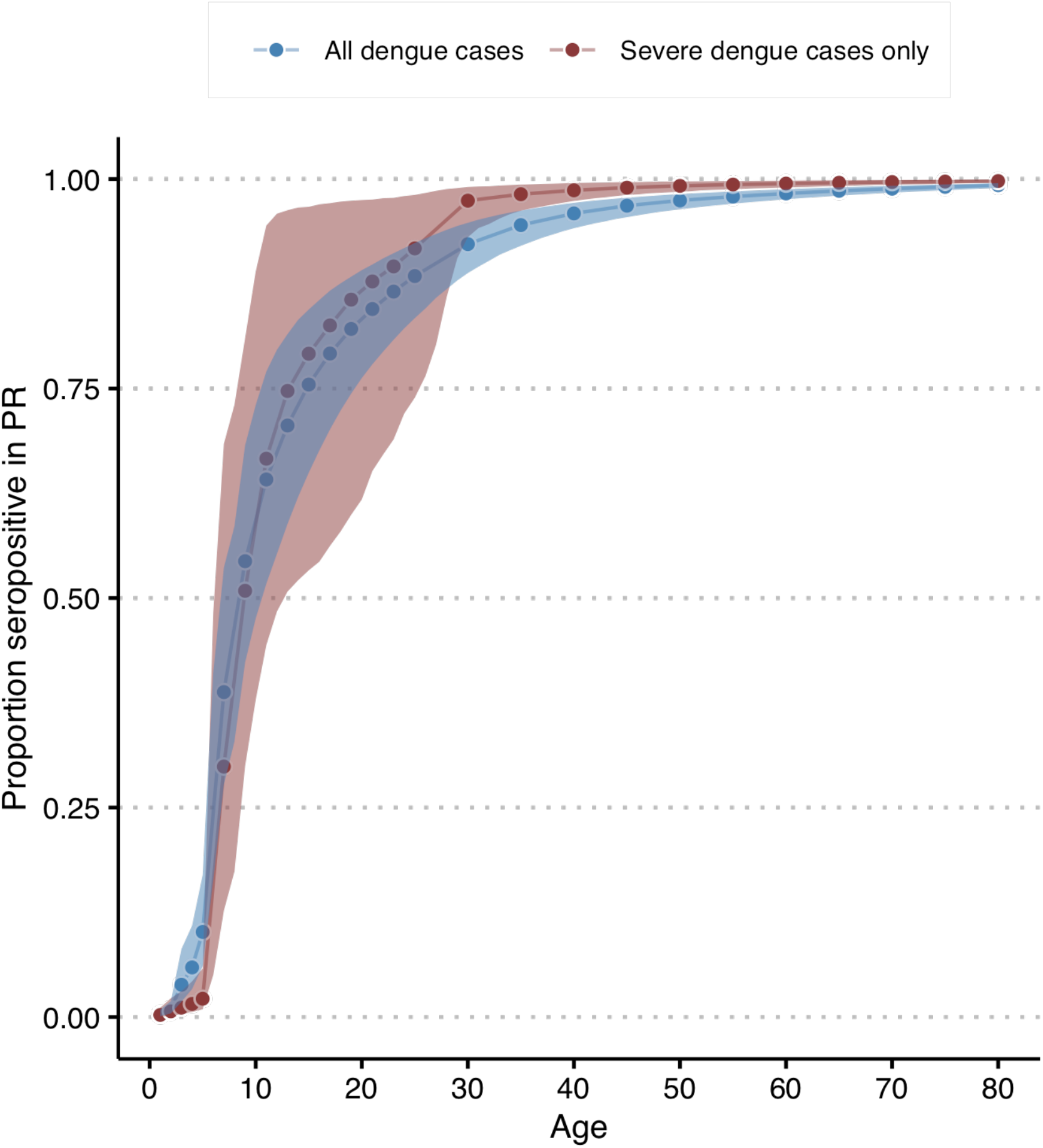
Age-specific seropositivity estimates for 2019 in Puerto Rico using all dengue cases and severe dengue cases only. Derived from model S where reported cases are secondary infections only. Shaded areas represent 95% confidence intervals.

While no cases were reported in American Samoa between 2010 and 2016, the model estimated low reporting in 2010–2016 (Figure 5) and high uncertainty for FOI prior to 2010 with the highest long-term average FOI of all locations at 1.4% (95% CI: 1.1–1.8%, Figure 4). The high estimated FOI in 2017–2018 translated to an estimated 2019 seroprevalence in 2019 for 5-year-olds of 46.5% (95% CI: 33.2– 61.6%, Figure 2B). Thus, the age structure of cases in 2017-2018, indicates that while this was a large outbreak, there was likely dengue transmission both in 2010-2016 and in preceding years. The FOI estimate for American Samoa was higher than the estimate for Puerto Rico, with similar estimated adult seroprevalence (91.9%, 95% CI: 84.8–95.7% for 35-year-olds, Figure 2B).

The US Virgin Islands had lower estimated long term average FOI (0.5%, 95% CI: 0.3–0.7%, Figure 4) and more stable estimated reporting (Figure 5). With a low FOI in recent years, the 5-year-old estimated seroprevalence for 2019 was low (5.1%, 95% CI: 2.3–11%, Figure 2C) but cumulative exposure in adults was still high, with 35-year-old estimated seroprevalence for 2019 of 86.1% (95% CI: 72.7–94.1%, Figure 2C).

Hawaii reported two outbreaks (2011 and 2014–2015) and Guam reported one (2019) during the study period (Figure 1) and both had less distinct age-specific case patterns (Figure 2D, 2F). The estimates for reporting largely reflected the reporting prior (Figure 4), indicating that there was little information between the case numbers and case age data to distinguish differences in reporting versus FOI. Nonetheless, long-term average FOI estimates were low for both jurisdictions (Hawaii: 0.001%, 95% CI: 0.0003–0.004%; Guam: 0.003%, 95% CI: 0.0008–0.013%; Figure 4), with significant increases in the years when the outbreaks were reported. The 2019 seroprevalence estimates were low for all age groups in both locations: 6.3% (95% CI: 3.9–12.3%, Figure 2D) and 2.6% (95% CI: 0.8–8.9%, Figure 2F) for 35-year-olds in Hawaii and Guam, respectively.

Florida was unique in that 90.5% of cases had reported travel histories. To assess local transmission within the state, we therefore only analyzed cases without reported travel (Figure 2E). Local dengue cases were reported in multiple years and had a distinctly different age profile for cases compared to other locations, with no reported cases in younger age groups and many among adults, which resulted in the largest uncertainty for seroprevalence estimates, especially for older age groups (Figure 2E). Considering all cases, the uncertainty for estimated seroprevalence in 35-year-olds was very high, with a 95% CI of 3.5% to 98.1%. Among local cases only, uncertainty was still high, but indicated more confidence in lower seroprevalence for younger adults, 9.6% (95% CI: 2.0–28.1%) for 35-year-olds. This lower seroprevalence estimate for local cases indicates a relatively low historical transmission intensity locally, implying that many imported secondary cases likely also had their primary infections outside of Florida. The long-term mean FOI for Florida considering only local cases was 0.001% (95% CI: 0.0002–0.008%). Similar to Hawaii and Guam, there was insufficient information to further resolve the probability of reporting a case and the estimated reporting probabilities resembled the prior.

Across the six locations, estimated long-term average FOI was highest for American Samoa and Puerto Rico, followed by the US Virgin Islands, with much lower estimates for Hawaii and Guam. We also assessed the average yearly FOI estimates for each location and found that for the higher FOI locations (Puerto Rico, American Samoa, and US Virgin Islands), the yearly average was lower for the period which included surveillance data (2010–2019) compared to estimates for time periods prior to 2010, for which ArboNET data are not available. In contrast, the estimated FOI for the 2010–2019 period were higher than the historical period for Hawaii and Guam.

## Discussion

Passive surveillance is the most common approach for dengue surveillance globally [30–32]. While passive surveillance can be highly effective for monitoring longitudinal changes in incidence, insight into disease burden is challenging as care seeking behavior, access to care, case definitions, reporting, and resources can differ substantially over time and across jurisdictions. Differences in these components can mean that actual increases, decreases, or jurisdictional differences in dengue burden are difficult to disentangle from reporting dynamics. The importance of understanding dengue burden beyond reported incidence has grown in recent years, with the advent of dengue vaccines which can be used to reduce the burden, but also require consideration of levels of preexisting population immunity for effective deployment [33]. Here, we extended a previously developed Bayesian model to leverage the age distribution among cases reported via passive surveillance in the US to estimate yearly DENV force of infection, yearly reporting probabilities, and seroprevalence for six jurisdictions with very different histories of reported dengue incidence.

Across the six jurisdictions, the estimated dengue burden was highly heterogeneous and FOI estimates did not appear be higher over time, except in Hawaii and Guam. We found evidence of high long-term transmission intensity in Puerto Rico, American Samoa, and US Virgin Islands (FOI and high seroprevalence in adults), despite different histories of dengue in each location. Hawaii, Florida, and Guam also differed in the frequency and magnitude of reported outbreaks, but all had considerably lower long-term FOIs.

The higher estimated FOI in Puerto Rico, American Samoa, and US Virgin Islands reflect different dynamics. Puerto Rico has consistently reported dengue cases for at least three decades ([34,35], timeline by [6]), while dengue reporting in American Samoa and the US Virgin Islands is more sporadic. Outbreaks have been reported in American Samoa in the 1970s, 1990s, 2000s, and 2017–2018 [36–38] and in the US Virgin Islands in the 1990s, 2000s, and in 2012–2013 [6,39]. Here, we found that despite these different apparent dynamics, the estimated long-term FOI and seroprevalence in these locations were similar, with a high likelihood that individuals experienced a secondary infection by the time they reached 10 years old (point estimates between 42% and 59%). Three seroprevalence studies support these findings. First, a study in the municipality of Patillas in the southeastern part of Puerto Rico [40] found a 42.5% seroprevalence in 10–11 year-olds in 2007–2008. For 10-year-olds, we estimated a seroprevalence of 57.2% (95% CI: 45.3–69.6%) in 2010 — the earliest estimate available for our study — in Puerto Rico. A 2010 seroprevalence study in American Samoa found a seroprevalence of 95.6% (95% CI 93.9–96.8%) for 18–87 year-olds, with slightly lower estimates for 18–25 year-olds (89.1%, 95% CI: 84.0–92.6%) [41]. For the same age groups and year, using only passive surveillance data, we estimated seroprevalence of 95.9% (95% CI: 86.9–98.5%) and 86.9% (95% CI: 63.6–95.6%) for these respective age groups. More recently, CDC’s updated traveler risk classifications re-categorized US Virgin Islands from “Sporadic/Uncertain” to “Frequent/Continuous” [42]. Additionally, a cross-sectional seroprevalence study conducted in US Virgin Island schools in April 2022 estimated a seroprevalence of 42% (95% CI: 26–60%) and 54% (95% CI: 18–89%) among 10 and 13 year-olds respectively [43], while our model estimated a seroprevalence of 34% (95% CI: 18–54%) and 41% (95% CI: 26–61%) in 7 and 13 year-olds for 2019, 3 years prior. No major outbreak has been reported in the US Virgin Islands between 2019 and 2022. The high level of immunity found in the US Virgin Islands with our model confirms that dengue transmission intensity in this territory is likely higher than previously suspected.

In Hawaii, low FOI and seroprevalence estimates are consistent with several possible histories: small intermittent outbreaks, rare large epidemics, or continuous low intensity transmission. No autochthonous cases had been reported in Hawaii between 1944 and 2001, but recent outbreaks were reported in 2001–2002 [44], 2011, and 2015–2016 [45]. Given some reporting issues during these outbreaks (see [46]), the seroprevalence profile in Hawaii should be interpreted with caution. Although the results here do not differentiate between the possible histories, they do indicate that substantial and sustained undetected transmission in recent decades is highly unlikely. Further serological data collection could help decipher between these hypotheses.

In Florida, we found a low FOI and a low level of immunity, suggesting low circulation of the virus. Frequent introductions of dengue viruses by travelers result in frequent opportunities for local transmission, so FOI estimates likely reflect both this relatively high introduction risk and limited local transmission. The travel and migration history of Florida residents may also contribute to a higher seroprevalence among older individuals than what would be expected from local transmission alone. This could explain the higher case numbers in older adults, high uncertainty in seroprevalence estimates for those adults, and the better fit of the secondary case model to the Florida data. More detailed data on histories of travel and dengue exposure could potentially provide more resolution on historical patterns of transmission in Florida as compared to exposures acquired elsewhere that impact the patterns of dengue seen in Florida today.

Our FOI and seroprevalence estimates in Guam were lower than for the other US territories with comparable environmental conditions. According to our model, primary infections, or a combination of primary and secondary infections dominated the 2019 outbreak in Guam, which is consistent with other studies that found no evidence of endemic circulation in Guam since World War II [38,47,48], potentially due to successful vector control strategies limiting dengue transmission on this territory [47].

Our model estimated time-varying reporting probabilities for all jurisdictions and found important variations across locations and years. In locations with lower long-term FOI estimates, there was little information in the case data to update the broad prior assumption for reporting probabilities of approximately 10–30%. However, for locations with higher long-term FOI, reporting was generally below 10% and showed substantial year-to-year variability. Other recent work indicates the possibility of similar variation between years [20,22] and particularly between epidemic and non-epidemic years [21], a pattern which we also found in the data and models analyzed here. These findings indicate the likely importance of accounting for variability in reporting probabilities over time.

We compared models accounting for reported cases as primary, secondary, or a combination of both primary and secondary infections, finding mixed evidence of fit across jurisdictions. For Florida, the secondary case model clearly fit better by leave-one-out metrics, while it was worse than the other models for Hawaii and Guam (Figure 3). However, detailed examination of fitted model estimates and log-likelihoods revealed that models for secondary infections provided better qualitative fits to the age profiles of cases in locations with higher seroprevalence and for years and age groups with higher incidence. Thus, despite equivocal out-of-sample performance, we found that the secondary-only model is likely preferrable in locations with high FOI or with high seroprevalence that may have been acquired elsewhere (Florida), while the primary or combined models may perform better in low FOI settings with limited previous exposure. For reporting, this finding implies that most cases reported in Puerto Rico, American Samoa, US Virgin Islands and Florida are secondary infections and the majority of reported cases in Hawaii and Guam are likely primary infections. This confirms that in endemic settings primary infections are largely unreported, but also suggests this pattern is slightly different in locations with more sporadic transmission. This spatial and temporal variation in reporting is likely to be a more general phenomenon.

While we were able to identify clear differences in transmission intensity across locations, this analysis provides limited insight into why those differences exist. Different levels of mobility and interactions with mosquitos between individuals within and between households likely lead to some differences. Environmental variables (e.g., temperature and rainfall) also impact the mosquito population and may drive differences in transmission intensity [49,50]. Furthermore, we accounted for previous infections as a condition for reporting and a condition for the force of infection. However, we did not consider how heterotypic and homologous immunity to different serotypes (or other flaviviruses) may substantially impact these dynamics.

Some limitations are worth noting. First, ArboNET includes dengue data since 2010 but in some jurisdictions few cases have been reported in this period, meaning that for some jurisdictions the models rely on relatively few data. While fitting the data in a Bayesian framework with informative priors reduces the possibility of misleading conclusions, it also means that high uncertainty remains around some important parameters (e.g., reporting in some jurisdictions). Heterogeneity in dengue exposure and disease across age and gender may result in both age-specific FOI and age-specific reporting probabilities. Neither of these possibilities are captured in our model implementation and would be difficult to assess without informative priors for that age-specific variation. In early model development, we found that to account for time-variable reporting and FOI, we needed to have informative priors for both components. Another possible area for advancing this type of model would be to further differentiate reporting probabilities for primary and secondary infections (see [14]). With informative priors, this could potentially provide further resolution on location specific differences in transmission where reported cases may be dominated by primary infections, secondary infections, or a mix.

Our study shows that analyzing age-specific case notification data with catalytic models can provide invaluable insight into dengue virus transmission dynamics beyond simple case counts, including estimates of transmission intensity over time and of evolving population-level exposure (as an ongoing proxy for seroprevalence). These components can help identify where interventions such as dengue vaccines should be prioritized in the US and elsewhere or what populations may be at higher risk of secondary infections and therefore severe disease. These models may provide an important tool for assessing and monitoring dengue transmission risk in many locations where age-specific surveillance data already exist.

## Data Availability

The ArboNET surveillance data is available to researchers upon request. Code used to run the current analysis are available in the GitHub repository, ...

https://github.com/sarahkada/dengue-catalytic-model

## Acknowledgments

This work was supported in part by an appointment to S.K. to the Research Participation Program at the Centers for Disease Control and Prevention, administered by the Oak Ridge Institute for Science and Education through an interagency agreement between the US Department of Energy and CDC. Authors would like to thank Aidsa Rivera for support over dataset requests.

## Disclaimer

The findings and conclusions in this report are those of the authors and do not necessarily represent the official position of the Centers for Disease Control and Prevention.

## Code availability

Code to reproduce our analyses is available on GitHub at https://github.com/sarahkada/dengue-catalytic-model.

## Supporting information Captions

**S1 Figure. Prior distribution of the reporting hyperprior**. Dashed lines represent median prior value, at 10%.

**S2 Figure. Age distribution of reported severe and non-severe dengue cases (colored bars) in Puerto Rico from 2010 to 2019**. The grey bars represent the age distribution of the population (US census, 2010).

**S3 Figure. Yearly age distribution of dengue reported cases in Puerto Rico (A), American Samoa (B), US Virgin Islands (USVI, C), Hawaii (D), Florida (E), and Guam (F)**. Scales may be adjusted for years with fewer dengue cases reported.

**S4 Figure. Yearly age distribution of locally-acquired and imported dengue reported cases in Florida**. Scales are adjusted for years with fewer dengue cases reported.

**S5 Figure. Comparison of log likelihood samples by age group and years of models 1-3 using Puerto Rico data. Comparison of log likelihood samples by age group and years of models Primary, Primary & Secondary, and Secondary using Puerto Rico data**. Models’ log-likelihood values may overlap. The log likelihood value in a model is a measure of goodness of fit. The higher the value (i.e., closer to 0), the better.

**S6 Figure. Heatmap comparison between models Primary, Primary and Secondary, and Secondary of log likelihood median samples and cases by age group and years using Puerto Rico data**. The log likelihood value in a model is a measure of goodness of fit. Here, models with log likelihood values closest to 0 were plotted.

**S7 Figure. Reported cases model fit (all cases and severe cases only) by age group in Puerto Rico from 2010 to 2019**. Points represent cases reported to ArboNET while lines represent model fit to Primary, Primary & Secondary, and Secondary models. Shaded areas represent the best model 95% CIs.

**S8 Figure. Reported severe cases model fit (severe cases only) by age group in Puerto Rico from 2010 to 2019**. Points represent cases reported to ArboNET while lines represent model fit to Primary, Primary & Secondary, and Secondary models.

Shaded areas represent the best model 95% CIs.

**S9 Figure. Reported cases model fit by age group in American Samoa from 2010 to 2019**. Points represent cases reported to ArboNET while lines represent model fit to Primary, Primary & Secondary, and Secondary models.

Shaded areas represent the best model 95% CIs.

**S10 Figure. Reported cases model fit by age group in US Virgin Islands from 2010 to 2019**. Points represent cases reported to ArboNET while lines represent model fit to Primary, Primary & Secondary, and Secondary models.

Shaded areas represent the best model 95% CIs.

**S11 Figure. Reported cases model fit by age group in Hawaii from 2010 to 2019**. Points represent cases reported to ArboNET while lines represent model fit to Primary, Primary & Secondary, and Secondary models. Shaded areas represent the best model 95% CIs.

**S12 Figure. Reported cases model fit by age group in Florida from 2010 to 2019**. Points represent cases reported to ArboNET while lines represent model fit to Primary, Primary & Secondary, and Secondary models. Shaded areas represent the best model 95% CIs.

**S13 Figure. Reported cases model fit by age group in Guam from 2010 to 2019**. Points represent cases reported to ArboNET while lines represent model fit to Primary, Primary & Secondary, and Secondary models. Shaded areas represent the best model 95% CIs.

**S14 Alpha parameter posterior estimates with 95%CI (vertical bars) in Puerto Rico, Puerto Rico severe (using severe cases only), American Samoa, US Virgin Islands, Hawaii, Florida and Guam**.

**Table S1: Force of infection estimates in Puerto Rico, American Samoa, US Virgin Islands, Hawaii, Florida and Guam from 2010 to 2019**. Median and 95% Bayesian credible intervals.

